# Child Mortality in England after the Pandemic. Increasing Mortality and Inequalities

**DOI:** 10.1101/2024.05.24.24307855

**Authors:** David Odd, Sylvia Stoianova, Tom Williams, Peter Fleming, Karen Luyt

**Affiliations:** School of Medicine, Division of Population Medicine, Cardiff University, UK; National Child Mortality Database, Bristol Medical School, University of Bristol, St Michael’s Hospital, Southwell Street, Bristol, UK; Centre for Academic Child Health, Population Health Sciences, Bristol Medical School, University of Bristol, UK

**Keywords:** COVID-19, SARS-CoV-2, coronavirus, pandemic, mortality, death, child, infant

## Abstract

**Background:** During the COVID-19 pandemic children and young people (CYP) mortality in England reduced to the lowest on record, but it is unclear if the mechanisms which facilitated a reduction in mortality had a longer lasting impact, and what impact the pandemic, and its social restrictions, have had on deaths with longer latencies (e.g. malignancies). The aim of this analysis was to quantify the relative risk of childhood deaths, in England, before, during, and after the COVID pandemic and its social changes.

**Methods and Findings:** Mortality for each analysis year was calculated per 1,000,000 person years. Poisson regression was used to test for an overall trend across the time period, and tested if trends differed between April 2019 to March 2021 (Period 1)) and April 2021 to March 2023 (Period 2). This was then repeated for each category of death and demographic group. The underlying population profile was obtained from 2021 ONS Census data. 12,828 deaths were included in the analysis. 59.4% of deaths occurred under 1 year of age. Mortality rate (per 1,000,000 CYP per year) dropped from 272.2 (264.8-283.8) in 2019-20, to 242.2 (233.4-251.2) in 2020-21, increasing to 296.1 (286.3-306.1) in 2022-23. Overall, death rate reduced in Period 1 (RR 0.96 (0.92-0.99)) and then increased in Period 2 (RR 1.12 (1.08-1.16)). Asian (p<0.001), Black (p-0.012), and Other (p=0.003) ethnic groups, and those in more deprived areas (p<0.001), had evidence of an initial reduction mortality, and then a subsequent increase. Death rates for children from White (p=0.601) or Mixed (p=0.823) ethnic backgrounds, or those in the least deprived areas, did not change over the study period.

**Conclusions:** Different temporal profiles were seen across cause of death categories, with reassuring trends in deaths from Suicide and Intrapartum deaths (after pandemic peaks). However, for all other causes of death, rates are either static, or increasing. Overall child mortality in England is now higher than before the pandemic. In addition, any reductions in health inequalities seen moving into, and during, the pandemic have now disappeared.

## INTRODUCTION

Since the start of the COVID-19 pandemic there have likely been over 14 million excess deaths worldwide(1). However, the number of deaths from COVID in children and young people (CYP) was low; around 1% of CYP deaths during the pandemic were likely caused by the COVID-19 virus(2). In contrast, overall mortality for CYP reduced over, and around, the period of lockdown(3, 4), falling to the likely lowest on record; with dramatic reductions in infections and deaths from underlying conditions(5). By the end of March 2022, risk of death in England, for children, had returned to levels comparable to pre-pandemic rates(5), leaving a cohort of around 350 more children alive at that point due to previous reductions in mortality over the pandemic years. The reductions seen were similar across age groups, regions of England, sex and socio-economic deprivation; suggesting that community actions can reduce the social-economic patterning of childhood mortality in England(6). Since then, England, like most countries, has rescinded its COVID policies and legislation, and has mostly returned to pre-pandemic functioning(7). It is unclear if the public health mechanisms which helped reduce mortality across vulnerable groups has had a longer lasting impact, and what impact the pandemic, and its social restrictions, have had on deaths with longer latencies (e.g. malignancies), first developing during the highest period of disruption when later diagnosis, or treatment, may have occurred. Equally, while overall child mortality fell, concerns remained in some categories (e.g. suicide(8)) and urban communities, where lasting societal and behavioural changes may have occurred.

The aim of this analysis was to quantify the relative risk of childhood deaths, across the whole of England, before, during, and after the COVID pandemic and its social changes, and identify any changes that may have occurred, and the patterns of mortality, during this period.

## METHODS

### Data, Study Design and Population

The National Child Mortality Database (NCMD) commenced data collection on 1^st^ April 2019, and collects data from all Child Death Overview Panels (CDOPs) across England.(9) In England, all deaths of children (0-17 years old) are reviewed by CDOPs, and data collated by the NCMD, with death notifications required by statute within 48 hours. Data was downloaded on 14^th^ September 2023. The study population was derived from 2021 Office of National Statistics (ONS) census data(10), and includes measures of ethnicity, sex and age.

### Outcome

Deaths of all CYP, who died before their 18^th^ birthday, and had been born at, or over, 22 weeks of gestation, occurring from 1^st^ April 2019 until 31^st^ of March 2023 were identified. In this work, similar to our previous work(4), in order to obtain a provisional category of death, all child deaths reported to NCMD were coded by 3 independent coders (all paediatricians) to identify the likely category of the cause of death. All coders recorded a provisional category of death (see below) or that there was insufficient information provided. For each death, if two or more coders agreed on a category this was taken as the most likely category and where no two coders agreed, the category highest in the following hierarchy was used (based on categorisation used by CDOPs(11), in order of priority: Trauma, Malignancy, Underlying Medical Condition, Intrapartum event, Preterm Birth, Infection, SUDIC (Sudden Unexpected Death in Infant and Childhood).

### Covariates

In the initial notification to NCMD, the CDOPs report baseline characteristics of the child; from which the following data were derived: sex of individual (female, male, other (including not known)), age at death, ethnicity, and the child’s home postcode. The postcode was then mapped to a geographic measure, the Middle Layer Super Output Area (MSOA), which is derived at granularity of around 7200 people(12). This code was used to identify the CYP’s region of England, the rural/urban status, and a local measure of deprivation: the Index of Multiple Deprivation (IMD). The IMD is calculated using 7 main domains (income, employment, education, health, crime, access to housing and services, and living environment). The population split into 10 equal sized (by people) deciles 1 being most deprived and 10 least deprived(13). Ethnic Group was derived from NHS data, and is parent reported at source, and classified using ONS guidelines; Asian or Asian British (Bangladeshi, Chinese, Indian, Pakistani, Any other Asian background), Black or Black British (African, Caribbean, Any other Black background), Multi-racial (White and Asian, White and Black African, White and Black Caribbean, Any other Multi-racial background), White or White British (British, Irish, Any other White background, Gypsy or Irish traveller), and Other (Arab, Any other ethnic group).

### Statistical Analysis

Deaths of children occurring from 1^st^ April 2019 until 31^st^ March 2023, born at, or after 22 weeks of completed gestation, were identified and divided into four 12-month periods; April to March (2019-20, 2020-21, 2021-22 and 2022-23). Initially, the demographics for childhood deaths, split by the year (starting April 1^st^) were compared from reported data. Demographic comparisons were made using the Chi^2^ test. Rates of overall death, and for each category, were derived for each year (starting April 1^st^).

Next, the underlying population size and characteristics were derived from Office of National Statistics 2021 Census data. Number of deaths were compared across years using the Chi^2^ statistics, and risk of death, for each analysis year (April-March) was calculated per 1,000,000 children, with counts collapsed to number of events per month. A Poisson regression model was used to test for an overall trend across the 4 years, along with the relative risk of death (per year), using counts of death collapsed to number per month. The number of likely additional deaths seen, between April 2020 and March 2023, compared to the number predicted from reference year of 2019-20 data, was derived using a generalised linear model (Poisson family). This was repeated for each provisional category of death. Next, a linear spline model was fitted allowing different trends for April 2019 to March 2021 (Period 1) and April 2021 to March 2023 (Period 2); periods before and after the COVID pandemic’s peak). A p-value was derived to test for a change of trend at this mid-point. This was repeated for each cause of death and the child’s characteristics (above) and interaction p-values (if relevant) were derived as evidence that the profile may differ by the stratified characteristic (e.g. sex).

Finally, to quantify changes in health inequalities across the 4 years, the relative risk of death for children living in the more deprived half of England (Deciles 1-5), was compared to those in the less deprived half (Deciles 6-10), for each of the four years of data, alongside tests for trend across the two a-priori time periods (as above). This analysis was then repeated, comparing mortality for children from non-white ethnicities, and each specific ethnic group, with those from white backgrounds.

This work was reviewed by the Chair of the Central Bristol NHS Research Ethics Committee who confirmed that NHS ethical approval, including obtaining individual consent, was not required. Data are presented as number (%), risk per 100,000 person year (95% CI), relative risk ratio (95% CI), or excess deaths (number) with 95% CI. All tests were two-sided. Reporting follows the STROBE reporting guideline for observational studies. Analysis was performed using Stata Version 17.

### Patient and Public Involvement

Parent and public involvement guided the design and setting up of the NCMD at establishment and real-time child mortality surveillance system at the beginning of the COVID-19 pandemic.

## RESULTS

Over the 48 months, a total of 13,743 deaths were reported to NCMD, of which 915 were excluded for birth before 22 weeks gestation, leaving 12,828 deaths; 3229 deaths in 2019-20, 2852 in 2020-21, 3260 in 2021-22 and 3487 in 2022/23 (S1 Appendix). Over half (59.4% (7622/12,828) of deaths occurred under 1 year of age, 57.0% (7275/12,774) were male, 16.9% (2046/12,142) were of White ethnicity and London had more deaths than any other region (17.1% (2192/12,828).

Overall risk of death dropped from 274.2 (264.8-283.8) in 2019-20 to 242.2 (233.4-251.2) in 2020-21, then increased to 276.8 (267.4-286.5) in 2021-22, and then to 296.1 (286.3-306.1) in 2022-23 (p_trend_<0.001) (Table 1) (S2 Appendix). Across the whole time period, there was evidence of an increase in deaths from Preterm birth (p_trend_=0.046), Infection (p_trend_<0.001), Trauma (p_trend_<0.001), SUDIC (p_trend_<0.001) and Underlying Disease (p_trend_=0.006). There was little evidence for an overall trend in the risk of deaths from Malignancy (p_trend_=0.831), Intrapartum events (p_trend_=0.135), Substance abuse (p_trend_=0.363) or Suicide (p_trend_=0.849) over the study period. Over the 3 years since the pandemic started (April 2020 to March 2023), there were similar numbers of overall deaths (as predicted by the observed risk seen in 2019-20: -88 (-473 to 297)), but more deaths from Trauma (161 (70 to 252)) and SUDIC (149 (6 to 292)) and fewer from Underlying Disease (-312 (-534 to -90)).

**Table 1.** Risk (per 1,000,000 children) by year of death, stratified by cause of death.

| Measure | N | Pre-pandemic Period | Pandemic Period |  |  |  |
| --- | --- | --- | --- | --- | --- | --- |
|  |  | Risk (per 1,000,000 population) | Risk (per 1,000,000 population) |  |  | Excess deaths in 2020-2023 |
|  |  | 2019-2020 | 2020-2021 | 2021-2022 | 2022-2023 |  |
| All Deaths | 12,828 | 274.2 (264.8-283.8) | 242.2 (233.4-251.2) | 276.8 (267.4-286.5) | 296.1 (286.3-306.1) | -88 (-473 to 297) |
| Death by Cause |  |  |  |  |  |  |
| Malignancy | 1041 | 22.0 (19.4-24.8) | 22.4 (19.8-25.3) | 21.2 (18.7-24.0) | 22.8 (20.1-25.7) | 5 (-104 to 114) |
| Preterm Birth | 2860 | 59.3 (55.0-63.9) | 56.1 (51.9-60.6) | 64.3 (59.8-69.0) | 63.1 (58.6-67.8) | 64 (-116 to 244) |
| Intrapartum event | 686 | 14.2 (12.1-16.5) | 15.9 (13.7-18.3) | 15.7 (13.5-18.1) | 12.5 (10.5-14.7) | 18 (-70 to 106.2) |
| Infection | 643 | 14.1 (12.0-16.4) | 6.8 (5.4-8.5) | 13.6 (11.6-15.9) | 20.1 (17.6-22.9) | -21 (-108 to 66) |
| Trauma | 821 | 14.0 (12.0-16.3) | 16.6 (14.4-19.1) | 18.5 (16.1-21.1) | 20.5 (18.0-23.3) | 161 (70 to 252) |
| Substance Abuse | 55 | 1.7 (1.0-2.6) | 0.8 (0.3-1.6) | 0.8 (0.3-1.5) | 1.4 (0.8-2.2) | -25 (-54 to 4) |
| Suicide | 477 | 9.4 (7.8-11.4) | 10.2 (8.4-12.2) | 11.6 (9.7-13.7) | 9.3 (7.7-11.3) | 33 (-39 to 105) |
| SUDIC | 1881 | 36.8 (33.4-40.4) | 35.8 (33.4-40.4) | 42.3 (38.7-46.2) | 45.1 (41.3-49.1) | 149 (6 to 292) |
| Underlying Disease | 4056 | 92.7 (87.3-98.4) | 69.2 (64.5-74.1) | 84.7 (79.5-90.1) | 97.8 (92.2-103.6) | -312 (-534 to -90) |

Numbers are risks per 1,000,000 children per year

When investigating the difference between the period up-to the peak of the pandemic (March 2021) and that afterwards, there was good evidence that deaths reduced in the first period (RR 0.96 (0.92-0.99)) and then increased in the second (RR 1.12 (1.08-1.16)) (p_difference_<0.001) (Figure 1) (Table 2).

**Figure 1.**
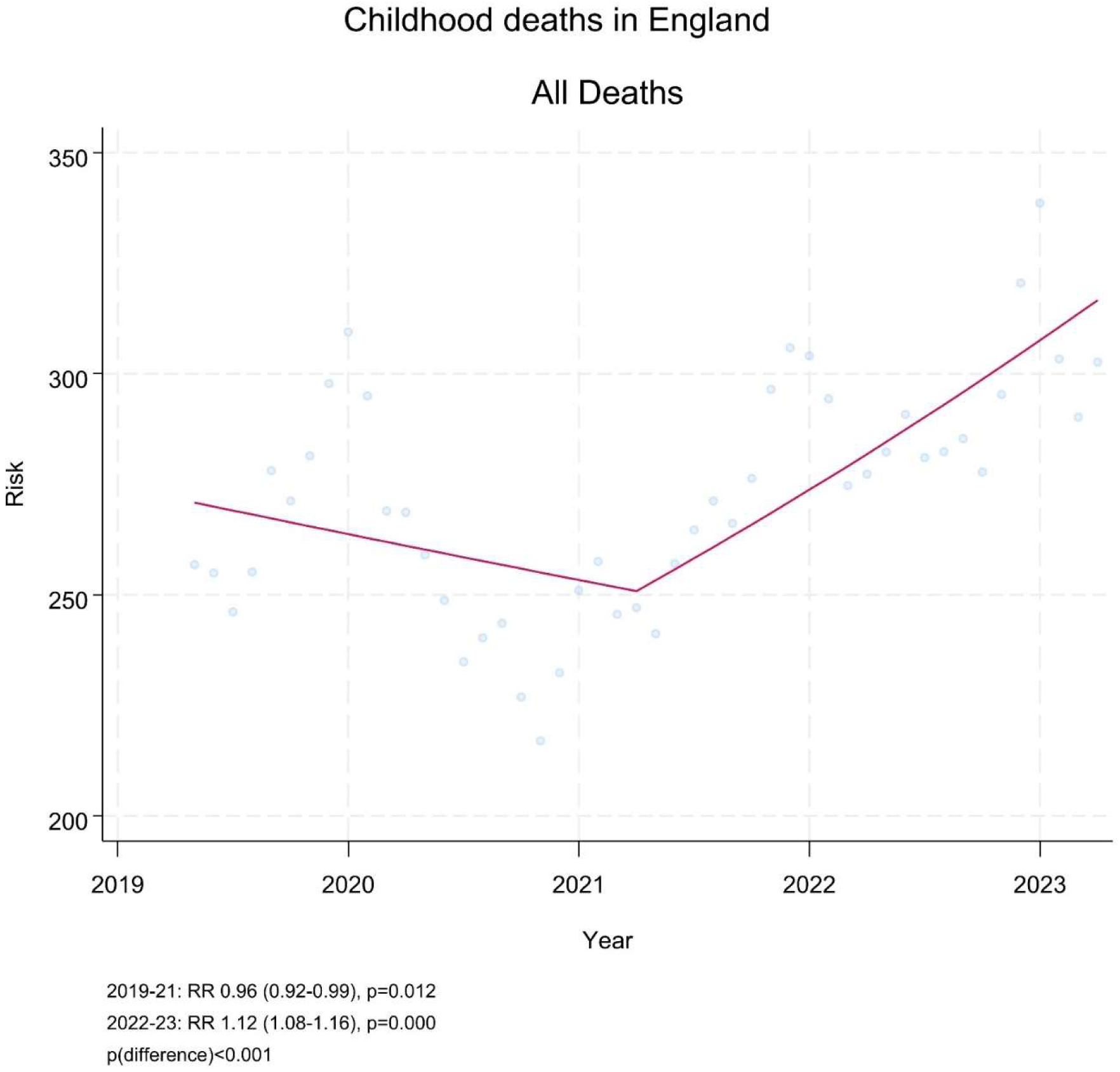
Risk of death per month (per 1,00,000 person years) and trends of risk across study period.

**Table 2.** Trends for overall, and categories of deaths, across the whole time period, and split between the two time periods.

| Measure | N | Trend over whole period (2019-2023) |  | Trend over 1<sup>st</sup> (2019-2021) or 2<sup>nd</sup> (2021-2023) period |  |  |
| --- | --- | --- | --- | --- | --- | --- |
|  |  | RR (95% CI) | p<sub>trend</sub> | Period 1<br>(April 2019-March 2021) | Period 2<br>(April 2021-March 2023) | Evidence of change in trajectory |
| All Deaths | 12,828 | 1.04 (1.02-1.05) | <0.001 | 0.96 (0.92-0.99) | 1.12 (1.08-1.16) | <0.001 |
| Death by Cause |  |  |  |  |  |  |
| Malignancy | 1041 | 1.01 (0.95-1.06) | 0.831 | 1.00 (0.89-1.13) | 1.01 (0.90-1.13) | 0.947 |
| Preterm Birth | 2860 | 1.03 (1.00-1.07) | 0.046 | 1.01 (0.94-1.09) | 1.05 (0.98-1.13) | 0.562 |
| Intrapartum event | 686 | 0.95 (0.89-1.02) | 0.135 | 1.15 (1.00-1.34) | 0.78 (0.68-0.91) | 0.004 |
| Infection | 643 | 1.21 (1.13-1.30) | <0.001 | 0.71 (0.60-0.84) | 1.80 (1.57-2.07) | <0.001 |
| Trauma | 821 | 1.12 (1.06-1.20) | <0.001 | 1.19 (1.03-1.38) | 1.07 (0.94-1.21) | 0.358 |
| Substance Abuse | 55 | 0.90 (0.71-1.13) | 0.363 | 0.52 (0.31-0.87) | 1.58 (0.94-2.67) | 0.021 |
| Suicide | 477 | 1.01 (0.93-1.09) | 0.849 | 1.17 (0.97-1.40) | 0.88 (0.74-1.04) | 0.073 |
| SUDIC | 1881 | 1.09 (1.04-1.13) | <0.001 | 1.04 (0.95-1.14) | 1.13 (1.04-1.23) | 0.281 |
| Underlying Disease | 4056 | 1.04 (1.01-1.07) | 0.006 | 0.85 (0.80-0.90) | 1.25 (1.18-1.32) | <0.001 |

Numbers are Relative Risk (95% CI)

**Table 3.** Relative risk of death, per year, over the whole 4 years, and split into the two periods; results stratified by CYP characteristics.

| Measure | N | Trend over whole period (2019-2023) |  | Trend over 1<sup>st</sup> (2019-2021) or 2<sup>nd</sup> (2021-2023) period |  |  |  |
| --- | --- | --- | --- | --- | --- | --- | --- |
|  |  | RR (95% CI) | p<sub>trend</sub> | Period 1<br>(April 2019-March 2021) | Period 2<br>(April 2021-March 2023) | Evidence of change<br>in trajectory | p<sub>interaction</sub> |
| Age of Death | 12,828 |  |  |  |  |  | <0.001 |
| <1 years | 7622 | 1.01 (0.99-1.03) | 0.244 | 0.97 (0.93-1.01) | 1.05 (1.01-1.10) | 0.041 |  |
| 1-4 years | 1475 | 1.07 (1.02-1.12) | 0.004 | 0.85 (0.76-0.94) | 1.31 (1.19-1.43) | <0.001 |  |
| 5-15 Years | 2643 | 1.09 (1.06-1.13) | <0.001 | 0.95 (0.88-1.03) | 1.23 (1.15-1.32) | <0.001 |  |
| 16-17 Years | 1088 | 1.08 (1.03-1.14) | 0.004 | 1.05 (0.93-1.18) | 1.11 (0.99-1.24) | 0.592 |  |
| Sex | 12,774 |  |  |  |  |  | 0.098 |
| Female | 5499 | 1.02 (1.00-1.05) | 0.082 | 0.97 (0.93-1.02) | 1.13 (1.09-1.18) | <0.001 |  |
| Male | 7275 | 1.06 (1.04-1.08) | <0.001 | 0.94 (0.89-0.99) | 1.10 (1.05-1.16) | <0.001 |  |
| Area of residence | 12,782 |  |  |  |  |  | 0.176 |
| Rural | 1450 | 0.91 (0.82-1.01) | 0.083 | 0.91 (0.82-1.01) | 1.15 (1.05-1.27) | 0.010 |  |
| Urban (London) | 2187 | 1.01 (0.97-1.05) | 0.657 | 0.93 (0.86-1.01) | 1.08 (1.00-1.17) | 0.041 |  |
| Urban (Not London) | 9145 | 1.05 (1.03-1.07) | <0.001 | 0.97 (0.93-1.02) | 1.12 (1.08-1.16) | <0.001 |  |
| Ethnicity | 12,142 |  |  |  |  |  | <0.001 |
| Asian or British Asian | 2242 | 1.10 (1.06-1.15) | <0.001 | 0.93 (0.86-1.01) | 1.28 (1.18-1.38) | <0.001 |  |
| Black or British Black | 1039 | 1.12 (1.06-1.18) | <0.001 | 0.97 (0.85-1.10) | 1.27 (1.14-1.42) | 0.012 |  |
| Mixed | 742 | 1.05 (0.99-1.12) | 0.097 | 1.04 (0.90-1.20) | 1.07 (0.94-1.22) | 0.823 |  |
| Other<sup>a</sup> | 361 | 1.13 (1.03-1.24) | 0.008 | 0.84 (0.68-1.04) | 1.45 (1.20-1.75) | 0.003 |  |
| White | 7758 | 1.05 (1.03-1.07) | <0.001 | 1.04 (0.99-1.08) | 1.06 (1.01-1.10) | 0.601 |  |
| Region of CDOP | 12,828 |  |  |  |  |  | 0.256 |
| East Midlands | 1054 | 1.03 (0.98-1.09) | 0.201 | 0.99 (0.88-1.12) | 1.07 (0.96-1.20) | 0.465 |  |
| East of England | 1233 | 1.02 (0.98-1.08) | 0.337 | 0.93 (0.83-1.04) | 1.12 (1.01-1.25) | 0.050 |  |
| London | 2192 | 1.01 (0.97-1.04) | 0.690 | 0.93 (0.85-1.01) | 1.09 (1.01-1.18) | 0.032 |  |
| North East | 633 | 1.11 (1.03-1.18) | 0.004 | 1.04 (0.88-1.22) | 1.17 (1.01-1.35) | 0.389 |  |
| North West | 1903 | 1.07 (1.03-1.11) | 0.001 | 0.96 (0.88-1.06) | 1.17 (1.08-1.27) | 0.015 |  |
| South East | 1706 | 1.03 (0.99-1.07) | 0.191 | 0.95 (0.86-1.04) | 1.11 (1.01-1.21) | 0.069 |  |
| South West | 947 | 1.01 (0.96-1.07) | 0.644 | 0.97 (0.85-1.10) | 1.05 (0.94-1.19) | 0.459 |  |
| West Midlands | 1721 | 1.06 (1.02-1.11) | 0.005 | 0.97 (0.88-1.07) | 1.15 (1.05-1.25) | 0.002 |  |
| Yorkshire and the Humber | 1439 | 1.07 (1.02-1.12) | 0.003 | 0.97 (0.88-1.08) | 1.16 (1.06-1.26) | 0.052 |  |
| Deprivation Decile | 12,753 |  |  |  |  |  | <0.001 |
| 1/2 (Most deprived) | 4377 | 1.06 (1.04-1.09) | <0.001 | 0.95 (0.89-1.01) | 1.18 (1.12-1.25) | <0.001 |  |
| 3/4 | 2904 | 1.04 (1.01-1.08) | 0.014 | 0.97 (0.90-1.04) | 1.11 (1.04-1.19) | 0.002 |  |
| 5/6 | 2236 | 1.00 (0.97-1.04) | 0.905 | 0.92 (0.85-1.00) | 1.08 (1.00-1.17) | 0.033 |  |
| 7/8 | 1760 | 1.02 (0.98-1.07) | 0.274 | 0.97 (0.88-1.07) | 1.07 (0.98-1.17) | 0.233 |  |
| 9/10 (Least deprived) | 1476 | 1.06 (1.01-1.10) | 0.014 | 1.05 (0.95-1.16) | 1.07 (0.97-1.17) | 0.832 |  |

(a) Other ethnic group (Arab, Any other ethnic group)
Numbers are Relative Risk (95% CI)

### Relative Risks are per year with 95% confidence intervals. Risk per month smoothed over 5 month period

A similar profile was seen for deaths from underlying disease, from infection, and from substance abuse across the time period (Infection, p<0.001; Underlying Disease, p<0.001; Substance Abuse, p=0.021) (Figure 2). In contrast, for deaths after Intrapartum events, an increasing risk in the first period (RR) was followed by a reduction in the second (RR) (p=0.004); and a similar, if weaker, pattern was seen with deaths after suicide (p=0.073).

**Figure 2.**
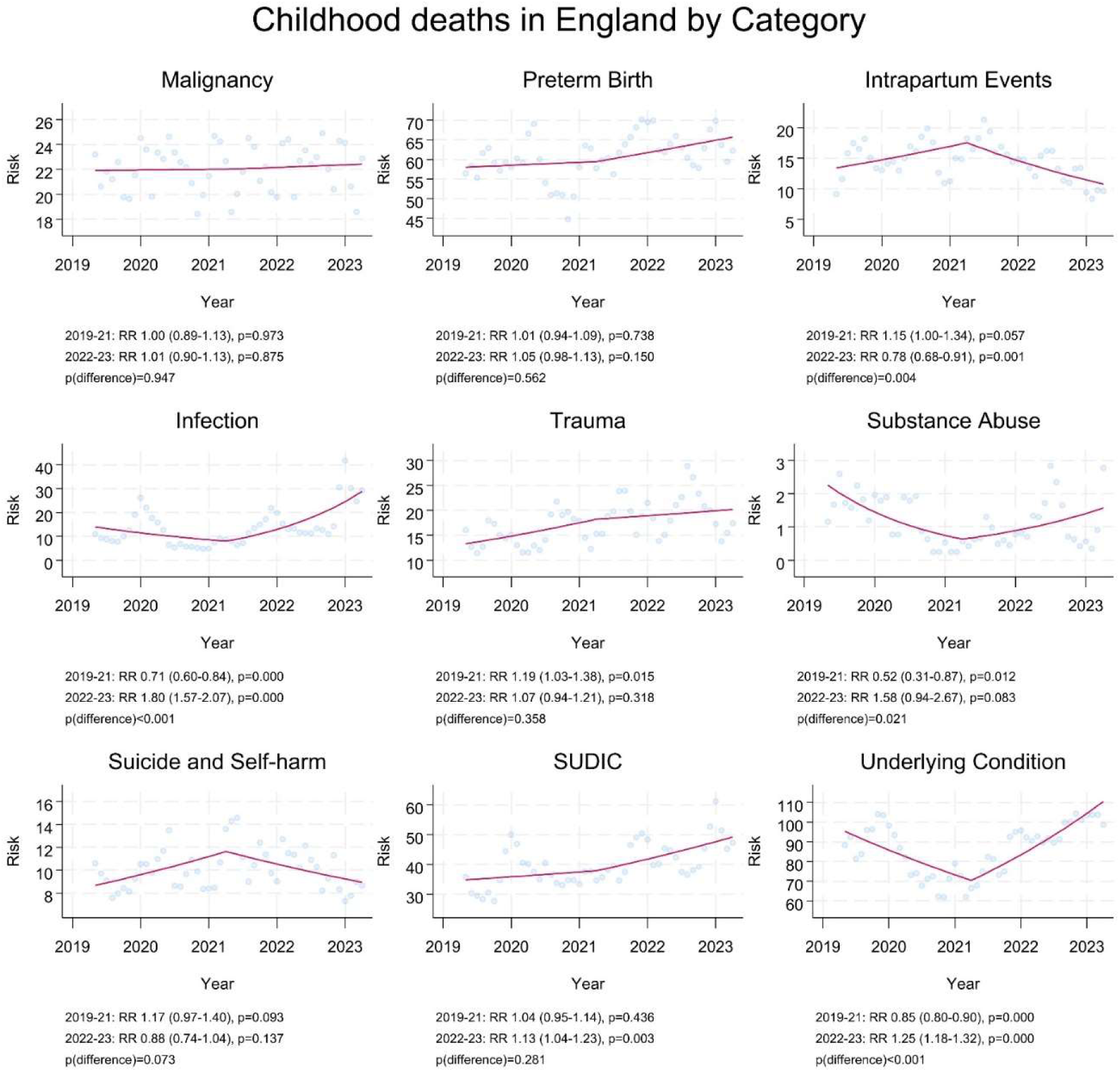
Risk of death per month (per 1,00,000 person years) and trends of risk across study period, split by category of death

Deaths from preterm birth, trauma and SUDIC, had increases in risk across the entire 4 year study period, with little evidence of a change in trajectory during the pandemic (Preterm birth, p=0.562; Trauma, p=0.358; SUDIC, p=0.281). There was no change in the risk of death from Malignancy overall (p=0.831), or any change in trend during the pandemic (p=0.947).

### Relative Risks are per year with 95% confidence intervals. Risk per month smoothed over 5 month period

Repeating the analysis, split by child characteristics, suggested that patterns of changing mortality were not modified by sex (p=0.098), area of residence (p=0.176) or region of England (p=0.256). However, the profile did differ by age (p<0.001), with mortality clearly dropping and then rising for children between 1 and 4 years old (p<0.001). A similar pattern was seen in children under 1, and between 5 and 15 years, although evidence of an initial drop was weaker in both groups (Infants RR 0.97 (0.93-1.01); 5-15 years RR 0.95 (0.93-1.18)). The risk appeared to increase across the whole time period for children in the oldest age group (16 and 17 years old) (RR 1.08 (1.03-1.14)). The pattern of mortality also appeared to differ by ethnicity with children from Asian or Asian British (p<0.001), Black or Black British (p=0.012) and Other (p=0.003) groups having a measurable reduction in mortality across the first period, and then an increase in the second. For children from White and Mixed background, any possible trend did not appear to be affected by the pandemic, with a general risk increase across the 4 years for white CYP (RR 1.05 (1.03-1.07)) and only weak evidence of an increase for those from a Mixed background (RR 1.05 (0.99-1.12)). For measures of deprivation, there was evidence of a change in risk trajectory for the 3 most deprived quintiles across the pandemic, with a decrease in the first half, and rise in the second. Children in the 2 least deprived quintiles had similar trends throughout the 4 years, with a general increase for the least deprived group (Deciles 9/10, RR 1.06 (1.01-1.10), or little evidence of change (Deciles 7/8, RR 1.02 (0.98-1.07)).

Finally, the relative risk of dying in the more deprived half of England remained similar across the first two years of this study (p_trend_=0.526) (Figure 3, S3 Appendix), but there was weak evidence that inequalities increased in the second half (p_trend_=0.093), with the highest RR seen in the final year (2022-23; RR 2.47 (2.30-2.66)). The relative risk of dying in children from Non-white backgrounds, compared to White children, reduced over the 1^st^ period (p_trend_=0.021), but then increased (p_trend_<0.001), to, again, the highest RR seen across the 4 years (2022-23; RR 1.70 (1.59-1.83).

**Figure 3.**
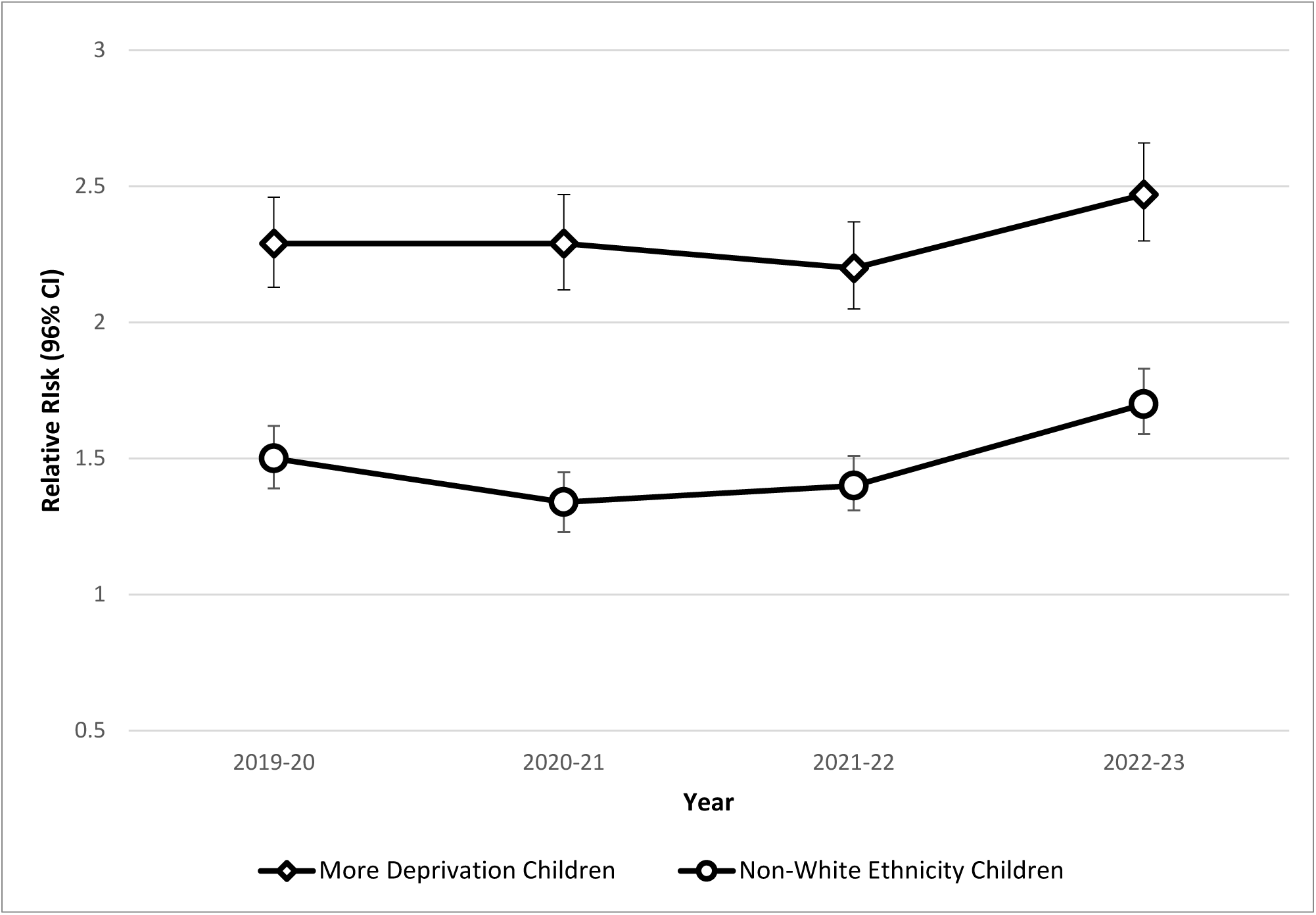
Relative Risk of childhood death for children in the more deprived (compared to the less deprived) areas, and children of non-white ethnicities (compared to those from white background), by year

## DISCUSSION

While the number of deaths in children and young people (CYP) in England overall dropped during the first year of the COVID-19 pandemic, any reduction of mortality has now been lost by increased risks of death in the subsequent two years.

Overall, mortality since the start of the pandemic is now higher than the year preceding the pandemic, and this profile is seen across a number of causes of death; including Infections. In contrast some causes of death (notably preterm birth, SUDIC, and trauma) have continued to increase over the 4 years of the study period. Two categories (Suicide and Intrapartum events) may have increased over the pandemic, but risk is now reducing back to similar baseline levels. The rate of reduction in mortality, and its subsequent return to pre-pandemic levels appears to have been most striking in the most vulnerable CYP. Finally, overall increases in mortality appear highest in the oldest children. Indeed, the oldest group here (16/17 years old) demonstrate a steady increase in mortality across the 4 years studied.

### Limitations

This work is based on the ongoing data collection from the NCMD, and previous work has shown good validation and coverage(14). While we had some missing data on some demographics (e.g. ethnicity), data completeness was generally good, and the primary analysis of risks was based on the statutory reporting of deaths. We have used ONS data to derive the population at risk, and this was estimated at a mid-point of the study. Changes may have biased our estimates (e.g. significant, differential migration, during the pandemic), and consequently absolute risk in particular should be interpreted with caution. However, the population came from a recent National Census and numbers of deaths are relatively small; making changes in the denominators unlikely to change the conclusions here. Fortunately, child death in England is rare, particularly for causes such as suicide and substance abuse deaths, which may further limit interpretation. Finally, the category of death in this work is provisional, based on information available at 48 hours, and further CDOP investigations may modify this in some cases.

### In Context

Overall, the number of deaths of CYP appears to be increasing, and is now above the level seen pre-pandemic(3, 4, 15), although four patterns across the 4 years appeared to exist. Two categories of death appeared to follow (and likely drive) this profile: deaths from infection and underlying disease. Both metrics are likely driven by a reduction in circulating pathogens, and public health initiatives to shield vulnerable CYP. These deaths are often in vulnerable CYP, either due to underlying health problems, or in vulnerable social groups; and the transient reduction in deaths across the pandemic appeared to have benefited these individuals most, reducing health inequalities across England. However, any benefits, temporary resources or learning from the pandemic, which helped reduce deaths of these children, appear to have been lost. Changes in behavior (e.g. enhanced hand washing)(16) or wider, state-based enhanced social support, which benefited our most vulnerable families, appear to be not delivering their previous benefits. As well as medically vulnerable children, we know that underlying mortality is higher in those from most deprived areas(17), and for some minority ethnic groups(18, 19). The benefits to these groups appeared to be the largest seen in this work, but again, any benefit appears to have disappeared, with rising mortality in those groups where, we saw transient reductions in health inequities.

Two further profiles of mortality were seen in this work. For two conditions (suicide and intrapartum events) we saw an increase in the number during the pandemic, and then a reduction leaving it. For both of these categories, a lack of data prior to 2019-20 limits the interpretation, and the trajectory of these deaths, had the pandemic not occurred, is unclear. We know suicide in CYP has risen in many countries over the last 10 years(20), and the apparent peak during lockdown in this work may represent the existing trend, or an effect of the social restrictions(21). However, the subsequent reduction was unexpected and further close monitoring is essential. For intrapartum events, the increase in deaths was unexpected, on a background of national initiatives to reduce brain injury, and death, around birth (e.g. the Each Baby Counts program)(22). Clearly, the subsequent reduction back to pre-pandemic levels is welcome, and may represent a lag in these initiatives embedding in practice. Perhaps the most surprising findings here were the conditions where, despite COVID, lockdown and broad societal change, little impact of the pandemic was apparent. Apparently indifferent to the pandemic impacts, we saw a general rise across the 4 years for deaths caused by preterm birth, trauma and SUDIC. Death by trauma is now the 4^th^ most common cause of childhood deaths in England, and while changes in practice around preterm birth(23); may influence some of the trend seen here, death after preterm birth continues to increase to the highest level since NCMD started collecting data, and remains a substantial contributor to the further widening of health inequalities in England(18, 24).

Finally, while malignancy, a common cause of childhood death, continues to show little, to no, overall influence from the pandemic, or generally over the time-period studied. Despite concerns related to later diagnosis or initiation of treatment, deaths from malignancy (the 3^rd^ highest category in this work) remains similar to 2019-20 levels, although we do know that this group was the one of the most vulnerable(2, 25). As with preterm birth, since the underpinning risk of death in this group is high(26) the effect of the pandemic may not have been considerable enough to be identified.

### Conclusion

Any mortality gains, often to vulnerable children, seen during the wide-ranging social changes put in place to reduce the spread and impact of COVID-19, have now dissipated. Overall child mortality in 2022-23 in England is now higher than before the pandemic. Three causes of death, trauma, SUDIC and preterm birth, appear to have continued to increase in incidence over the 4 years, with only Suicide and Intrapartum event deaths showing evidence of a reducing trend (although both of which may have increased during the pandemic). For all other causes of death, rates are either static, or worsening. In addition, the reductions in health inequalities seen during the pandemic in the most deprived children in England, and in our highest risk minority ethnic groups, have now reversed, with higher measures of inequalities than before the pandemic, for most vulnerable groups.

## Data Availability

The data underlying the results presented in the study may be available on request to the corresponding author, and subject to approval by Healthcare Quality Improvement Partnership (HQIP).

## Acknowledgements

We thank all Child Death Overview Panels (CDOPs) who submitted data for the purposes of this report and all child death review professionals for submitting data and providing additional information when requested.

Parent and public involvement is at the heart of the NCMD programme. We are indebted to Charlotte Bevan (Sands - Stillbirth and Neonatal Death Charity), Ann Chalmers (Child Bereavement UK) and Jenny Ward (Lullaby Trust), who represent bereaved families on the NCMD programme steering group.

We thank the NCMD team for technical and administrative support.

David Odd had full access to all the data in the study and takes responsibility for the integrity of the data and the accuracy of the data analysis.

## Ethics approval and consent to participate

The NCMD legal basis to collect confidential and personal level data under the Common Law Duty of Confidentiality has been established through the Children Act 2004 Sections M - N, Working Together to Safeguard Children 2018 (https://consult.education.gov.uk/child-protection-safeguarding-and-family-law/working-together-to-safeguard-children-revisions-t/supporting_documents/Working%20Together%20to%20Safeguard%20Children.pdf) and associated Child Death Review Statutory & Operational Guidance https://assets.publishing.service.gov.uk/government/uploads/system/uploads/attachment_data/file/859302/child-death-review-statutory-and-operational-guidance-england.pdf).

The NCMD legal basis to collect personal data under the General Data Protection Regulation (GDPR) without consent is defined by GDPR Article 6 (e) Public task and 9 (h) Health or social care (with a basis in law).

## Authors Contributions

DO: I declare that I participated in the study concept and design, contributed to acquisition, analysis and interpretation of data, drafting and review of the manuscript and that I have seen and approved the final version. SS: I declare that I participated in the study design, contributed to data acquisition, linkage, analysis and interpretation of analysis, drafting and review of the manuscript; and that I have seen and approved the final version.

TW: I declare that I participated in the study design, contributed to data acquisition, linkage, analysis and interpretation of data analyses, reviewing the manuscript; and that I have seen and approved the final version.

PF: I declare that I contributed to study design, interpretation of data analysis, reviewing the manuscript; and that I have seen and approved the final version.

KL: I declare that I obtained funding for this work, participated in the study concept and design, contributed to data acquisition and interpretation of data, drafting and reviewing the manuscript; and that I have seen and approved the final version.

## Transparency statement

Prof Karen Luyt (the manuscript’s guarantor) affirms that the manuscript is an honest, accurate, and transparent account of the study being reported; that no important aspects of the study have been omitted; and that any discrepancies from the study as originally planned have been explained.

## Availability of data

Aggregate data may be available on request to the corresponding author, and subject to approval by HQIP.

---

**S1 Appendix.**
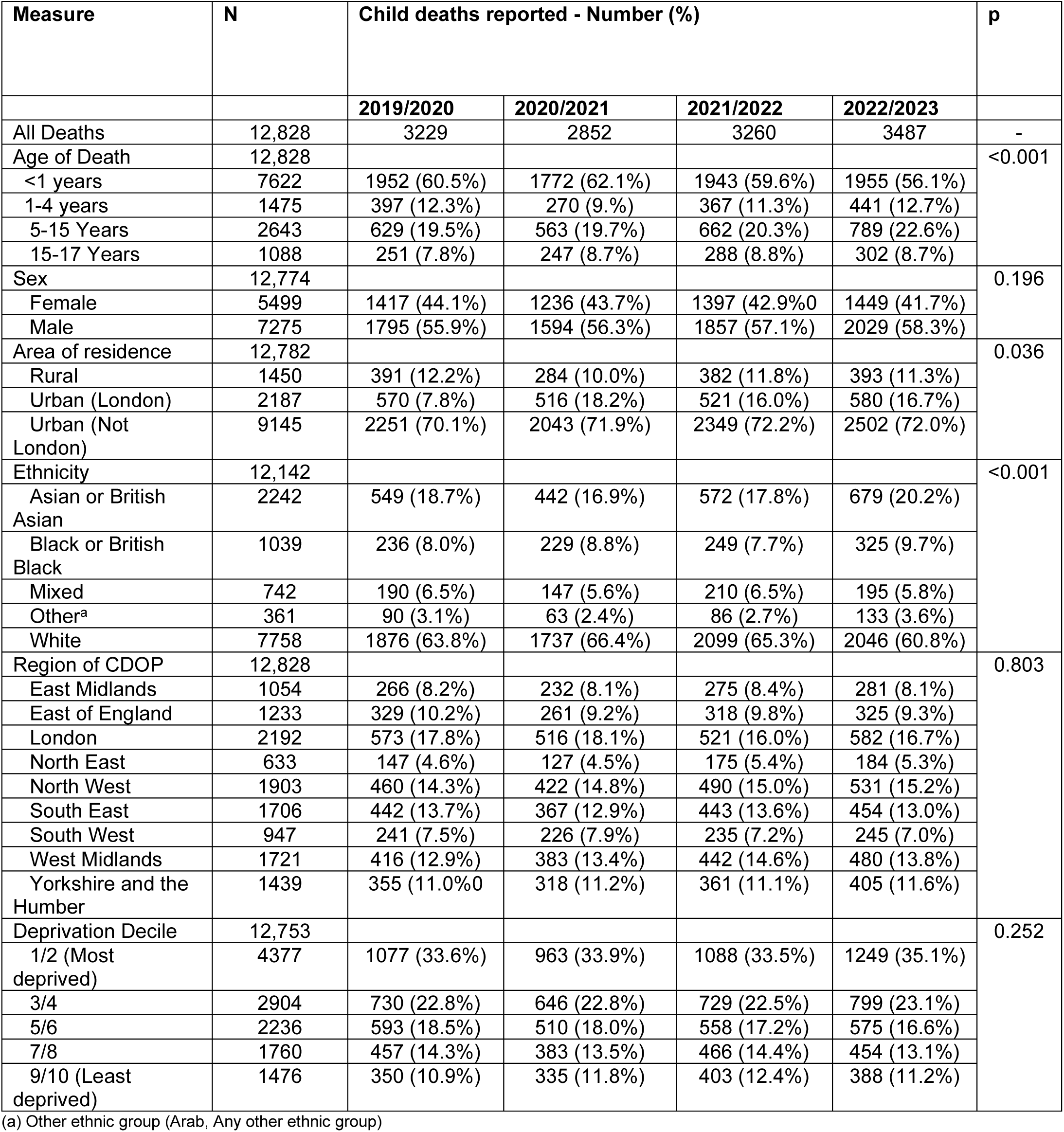
Characteristics of the populations of child deaths reported to NCMD in England between April 2019 and March 2023.

**S2 Appendix.** Causes of child deaths reported to NCMD in England between April 2019 and March 2023.

| Measure | N | Child deaths reported – Number (Percent) |  |  |  | p |
| --- | --- | --- | --- | --- | --- | --- |
|  |  | 2019/2020 | 2020/2021 | 2021/2022 | 2022/2023 |  |
| All Deaths | 12,828 | 3229 | 2852 | 3260 | 3487 | - |
| Death by Cause | 12,520 |  |  |  |  | <0.001 |
| Malignancy | 1041 | 259 (22.5%) | 264 (9.6%) | 250 (7.8%) | 268 (7.8%) |  |
| Preterm Birth | 743 | 699 (22.5%) | 661 (24.0%) | 757 (23.6%) | 743 (21.6%) |  |
| Intrapartum Event | 686 | 167 (5.4%) | 187 (6.8%) | 185 (5.8%) | 147 (4.3%) |  |
| Infection | 643 | 166 (5.3%) | 80 (2.9%) | 160 (5.0%) | 237 (6.9%) |  |
| Trauma | 821 | 165 (5.3%) | 196 (7.1%) | 218 (6.8%) | 242 (7.0%) |  |
| Substance Abuse | 55 | 20 (0.6%) | 10 (0.4%) | 9 (0.3%) | 16 (0.5%) |  |
| Suicide | 477 | 111 (3.6%) | 120 (4.4%) | 136 (4.2%) | 110 (3.2%) |  |
| SUDIC | 1881 | 433 (13.9%) | 419 (15.2%) | 498 (15.5%) | 531 (15.4%) |  |
| Underlying Disease | 4056 | 1092 (35.1%) | 815 (29.6%) | 997 (31.1%) | 1152 (33.4%) |  |

**S3 Appendix.** Relative risk of death, for each year of the study, compared by measures of deprivation and ethnicity.

| Measure | N | Year |  |  |  | Period 1<br>(April 2019-March<br>2021) | Period 2<br>(April 2021-March 2023) |
| --- | --- | --- | --- | --- | --- | --- | --- |
|  |  | 2019-2020 | 2020-2021 | 2021-2022 | 2022-2023 | p<sub>trend</sub> | p<sub>trend</sub> |
| Deprivation |  |  |  |  |  |  |  |
| Least Deprived Areas | 4307 | 1 (Ref) | 1 (Ref) | 1 (Ref) | 1 (Ref) |  | - |
| Most deprived areas | 8449 | 2.29 (2.13-2.46) | 2.29 (2.12-2.47) | 2.20 (2.05-2.37) | 2.47 (2.30-2.66) | 0.526 | 0.093 |
| Ethnicity |  |  |  |  |  |  |  |
| White | 7758 | 1 (Ref) | 1 (Ref) | 1 (Ref) | 1 (Ref) |  |  |
| Any non-White Ethnicity | 4384 | 1.50 (1.39-1.62) | 1.34 (1.23-1.45) | 1.40 (1.31-1.51) | 1.70 (1.59-1.83) | 0.021 | <0.001 |
| Asian or British Asian | 2242 | 1.72 (1.57-1.90) | 1.50 (1.35-1.66) | 1.61 (1.46-1.76) | 1.95 (1.79-2.13) | 0.029 | <0.001 |
| Black or British Black | 1039 | 1.60 (1.40-1.83) | 1.68 (1.46-1.92) | 1.51 (1.32-1.72) | 2.02 (1.79-2.27) | 0.297 | 0.002 |
| Mixed | 742 | 1.08 (0.93-1.25) | 0.90 (0.76-1.07) | 1.07 (0.93-1.23) | 1.02 (0.88-1.18) | 0.998 | 0.908 |
| Other<sup>a</sup> | 361 | 1.31 (1.06-1.62) | 0.99 (0.77-1.27) | 1.12 (0.90-1.39) | 1.63 (1.36-1.95) | 0.055 | 0.001 |

(a) Other ethnic group (Arab, Any other ethnic group)
Numbers are Relative Risk (95% CI)

## REFERENCES

1. 14.9 million excess deaths associated with the COVID-19 pandemic in 2020 and 2021. World Health Organisation. 2022.

2. Odd D, Stoianova S, Williams T, Thursby-Pelham A, Ladhani S, Oligbu G, et al. Deaths in Children and Young People in England following SARS-CoV-2 infection during the first two years of the pandemic: a national study using linked mandatory child death reporting data. Research Square. 2023.

3. Odd D, Stoianova S, Williams T, Fleming P, Luyt K. Child mortality in England during the first year of the COVID-19 pandemic. Arch Dis Child. 2022;107(3):e22.

4. Odd D, Stoianova S, Williams T, Sleap V, Blair P, Fleming P, et al. Child mortality in England during the COVID-19 pandemic. Arch Dis Child. 2022;107(1):14–20.

5. Odd D, Stoianova S, Williams T, Fleming P, Luyt K. Child Mortality in England During the First 2 Years of the COVID-19 Pandemic. JAMA Netw Open. 2023;6(1):e2249191.

6. Odd D, Stoianova S, Sleap V, Williams T, Cook N, McGeehan L, et al. Child Mortality and Social Deprivation. National Child Mortality Database (UK). 2021.

7. Wise PH, Chamberlain LJ. Adversity and Opportunity—The Pandemic’s Paradoxical Effect on Child Health and Well-being. JAMA Pediatrics. 2022;176(7):e220063-e.

8. Odd D, Williams T, Appleby L, Gunnell D, Luyt K. Child suicide rates during the COVID-19 pandemic in England. J Affect Disord Rep. 2021;6:100273.

9. Child Death Review: Statutory and Operational Guidance (England). HM Government (UK). London; 2018.

10. Census 2021: Office for National Statistics (ONS); 2022 [Available from: https://www.ons.gov.uk/census.

11. Sidebotham P, Fox J, Horwath J, Powell C, Shahid P. Preventing Childhood Deaths. Department for children, schools and families (UK). 2008.

12. Area Classifications in Great Britain.

13. McLennan D, Noble S, Noble M, Plunkett E, Wright G, Gutacker N. The English Indices of Deprivation 2019: Technical Report. Ministry of Housing, Communities and Local Government. 2019.

14. Battersby C, Statnikov Y, Santhakumaran S, Gray D, Modi N, Costeloe K. The United Kingdom National Neonatal Research Database: A validation study. PloS one. 2018;13(8):e0201815.

15. Williams T, Sleap V, Stoianova S, Rossouw G, Cook N, Odd D, et al. NCMD second annual report. National Child Mortliaty Database (UK). 2021.

16. Sansone V, Miraglia Del Giudice G, Della Polla G, Angelillo IF. Impact of the COVID-19 pandemic on behavioral changes in healthcare workers in Italy. Front Public Health. 2024;12:1335953.

17. Odd D, Stoianova S, Williams T, Odd D, Kurinczuk JJ, Wolfe I, et al. What is the relationship between deprivation, modifiable factors and childhood deaths: a cohort study using the English National Child Mortality Database. BMJ Open. 2022;12(12):e066214.

18. Odd DE, Stoianova S, Williams T, Odd D, Edi-Osagie N, McClymont C, et al. Race and Ethnicity, Deprivation, and Infant Mortality in England, 2019-2022. JAMA Netw Open. 2024;7(2):e2355403.

19. Freemantle N, Wood J, Griffin C, Gill P, Calvert MJ, Shankar A, et al. What factors predict differences in infant and perinatal mortality in primary care trusts in England? A prognostic model. Bmj. 2009;339:b2892.

20. Padmanathan P, Bould H, Winstone L, Moran P, Gunnell D. Social media use, economic recession and income inequality in relation to trends in youth suicide in high-income countries: a time trends analysis. J Affect Disord. 2020;275:58–65.

21. Gunnell D, Appleby L, Arensman E, Hawton K, John A, Kapur N, et al. Suicide risk and prevention during the COVID-19 pandemic. The Lancet Psychiatry. 2020;7(6):468–71.

22. Each Baby Counts: 2019 Progress Report. Royal College of Obstetricians and Gynaecologists; 2020 2020.

23. Davis PJ, Fenton AC, Stutchfield CJ, Draper ES. Rising infant mortality rates in England and Wales. BMJ. 2018;361.

24. Odd D, Williams T, Stoianova S, Rossouw G, Fleming P, Luyt K. Newborn Health and Child Mortality Across England. JAMA Netw Open. 2023;6(10):e2338055.

25. Wilde H, Tomlinson C, Mateen BA, Selby D, Kanthimathinathan HK, Ramnarayan P, et al. Hospital admissions linked to SARS-CoV-2 infection in children and adolescents: cohort study of 3.2 million first ascertained infections in England. Bmj. 2023;382:e073639.

26. Odd D, WIlliams T, Stoianova S, Sleap V, Glover N, Rossouw G, et al. The Contribution of Newborn Health to Child Mortality across England.: National Child Mortality Database (UK); 2022 [Available from: https://www.ncmd.info/wp-content/uploads/2022/07/Perinatal-FINAL.pdf.

